# Genetic and early environmental predictors of adulthood self-reports of trauma

**DOI:** 10.1101/2021.06.09.21258603

**Authors:** Alicia J. Peel, Kirstin L. Purves, Jessie R. Baldwin, Gerome Breen, Jonathan R.I. Coleman, Jean-Baptiste Pingault, Megan Skelton, Abigail R. ter Kuile, Andrea Danese, Thalia C. Eley

## Abstract

**Background:** Evidence suggests that retrospective self-reports of childhood trauma are associated with a greater risk of psychopathology in adulthood than are prospective measures. However, it remains unclear why retrospectively reported trauma confers a greater risk for poor outcomes. Investigating the heritable characteristics and environmental adversities associated with measures of self-reported trauma could increase understanding of this risk pathway to psychopathology.

**Methods:** Our sample included 3,963 unrelated individuals from the Twins Early Development Study. We tested whether polygenic scores for 21 psychiatric, cognitive, anthropometric and personality traits were associated with childhood emotional and physical abuse retrospectively self-reported in adulthood. To assess the presence of gene-environment correlation, we investigated whether these associations remained after controlling for composite scores of environmental adversity between birth and age 16.

**Results:** Retrospectively self-reported childhood trauma was associated with polygenic scores for autism spectrum disorder (ASD), body mass index and risky behaviours. When composite scores of environmental adversity were included in one model, only associations with parent-reported environmental adversity in middle childhood, self-reported environmental adversity in early adolescence and the polygenic score for ASD remained significant.

**Conclusions:** Retrospective self-reports of childhood trauma are associated with heritable characteristics of the reporter. Genetic predisposition to ASD may increase liability to experiencing or interpreting events as traumatic. Associations between genetic predisposition for risky behaviour and high BMI with self-reported childhood trauma may be environmentally-mediated. Studies of the association between retrospectively self-reported childhood trauma and later life outcomes should consider that genetically-influenced reporter characteristics may confound associations, both directly and through gene-environment correlation.

## Introduction

Childhood trauma is a well-established risk factor for numerous psychological disorders, including depression ^1^, anxiety ^2^, psychosis ^3^, personality disorders ^4^ and behavioural disorders ^5^. The experience of childhood trauma can be assessed using both prospective and retrospective measures. Prospective measures collect data from the time of the trauma occurring, and typically include official records and parent reports collected during early life. In contrast, retrospective measures are largely self-reported and assess personal narratives about past experiences of trauma.

It was generally assumed that retrospective reports capture the same experiences of trauma that would be detected by prospective measures ^6^. However, retrospectively self-reported childhood trauma has shown poor agreement with prospective accounts, indicating that these measures capture largely distinct groups of individuals, with potentially different risk pathways to psychopathology ^7^. Furthermore, evidence suggests that retrospectively reported trauma is associated with a greater risk of psychopathology in adulthood than prospective measures ^8–10^. For example, one study found that among children with prospective court records of maltreatment, the associated risk of psychopathology was minimal in the absence of retrospective self-reports of childhood trauma ^8^. In contrast, the risk associated with retrospective self-reports was high, whether or not there were corresponding prospective records. Despite this, there is little understanding of why retrospectively reported trauma may confer a greater risk for poor outcomes. Investigating the genetic and environmental correlates of retrospective reports could increase understanding of this risk pathway to psychopathology and has the potential to highlight new mechanisms for targeted intervention ^6^.

The increased risk of psychopathology associated with retrospective self-reports compared to prospective measures of trauma could be attributed to several factors, which are unlikely to occur in isolation. First, it could indicate that the subjective experience of trauma by itself, as captured by retrospective self-reports, has a greater influence on the development of adult psychiatric disorders than the occurrence of the event ^8^. Second, retrospective self-reports may be more open to differences in the interpretation of experiences as traumatic, the likelihood of recalling these experiences as traumatic, and the willingness to report traumatic events. These differences in reporting may be influenced by personality traits, psychopathology or cognitive biases ^10^. In turn, these characteristics could reflect or affect risk of psychopathology. Third, systematic differences between retrospective and prospective measures, for example in the reporter or type of measure (i.e. questionnaire vs. official records) could cause them to detect different types of events. Compared to prospective measures, retrospective measures may capture more cases of less severe forms of maltreatment ^7^. In addition, because retrospective self-reports are influenced by an individual’s perception, they might capture additional experiences throughout childhood that shape such perceptions ^11^. Indicators of challenging early environments, for example, child temperament, early separation from mother, harsh punishment, maternal external locus of control and negative life events, are found to be associated with retrospectively self-reported childhood trauma but not official records ^12^. In turn, the accumulation of experiences detected through self-reports may affect risk of psychopathology. These features of retrospective reports may mean that they are more likely to be influenced by individual differences in the reporter. These individual differences could include both genetic risk and environmental experiences.

In accordance with this, evidence suggests that retrospective self-reports are partly under genetic influence ^13,14^. Similarly, reporter characteristics including personality traits, psychopathology and cognitive biases have also been found to have a heritable component ^15–17^. As such, the heritability of reporter characteristics could, in part, explain the genetic component identified for retrospectively self-reported trauma ^18–20^. Numerous twin studies indicate overlapping genetic influences on self-reported negative life events and reporter characteristics, including personality ^21,22^, cognitive abilities ^23^, and psychopathology ^24,25^. Similarly, genome-wide association studies of retrospective self-reported childhood trauma find there is overlap with the specific genetic variants associated with personality, cognitive, behavioural, anthropometric and psychopathological traits, and find substantial genetic correlations between these traits and self-reported childhood trauma ^14,20^. For example, self-reported childhood trauma showed significant genetic correlations with neuroticism, years of schooling, smoking, body mass index (BMI), autism spectrum disorder (ASD) and major depression ^14^. In addition to exploring genetic correlations, genome-wide association data can be used to create polygenic scores. Such scores capture an individual’s genetic predisposition to a specific trait. These are created by summing the genetic variants that an individual carries, weighted by the size of their effect on the trait as estimated from a genome-wide association study ^26^. In a recent study of female nurses, polygenic scores representing genetic liability for attention deficit hyperactivity disorder (ADHD), major depression, neuroticism, schizophrenia, bipolar disorder, and ASD were all associated with a higher likelihood of reporting childhood abuse in mid-life ^27^. These findings suggest that associations between retrospectively self-reported childhood trauma and psychopathology are partly confounded by genetic predisposition to these outcomes, which influence both the likelihood of reporting traumatic events and of developing symptoms of a disorder.

However, these methods do not take into account the effect of environmental risk. As well as acting through the individual directly, genetics also impact a person’s environment. This process, termed gene-environment correlation (rGE), occurs when genetic factors influence both an individual’s liability for a trait and the environments that they are exposed to ^28^. There are three mechanisms of rGE: passive, active and evocative. In early childhood, genetic influence on the environment occurs primarily through passive, and to a lesser extent evocative, rGE ^29^. Passive rGE describes the association between the genotype that a child inherits, and the rearing environment that the parents create. For example, as maternal mental illness is associated with increased risk of child maltreatment ^30^, children exposed to maltreatment in this context will also be at elevated genetic risk for psychopathology. Evocative rGE refers to an individual’s genetically-influenced behaviours instigating certain responses from others in their environment. For example, the genetic factors that influence childhood antisocial behaviour have been found to be largely overlapping with those that influence their experience of physical discipline ^31^. As a child grows and interacts with their surroundings more, active rGE also becomes increasingly relevant. Active rGE describes the association between an individual’s genetically-influenced traits and the environments that they seek out. For example, genetic influences on affiliation with deviant peers have been identified, and are found to increase throughout adolescence to age 21 ^32^. This highlights how rGE effects increase as children and adolescents gain more control of their environments, resulting in a greater loading of genetic influence on those environments. While the influence of evocative rGE is proposed to persist across development, the influence of passive rGE is proposed to decline as it is replaced by more active forms from infancy to adolescence ^29^. This mechanism is anticipated to underlie the increasing heritability observed for psychopathological traits, such as depression, across development ^24^. These changes in the extent and nature of rGE throughout childhood highlight the importance of investigating these processes from a developmental perspective.

In order to understand the risk pathways through which retrospective reports of childhood trauma influence outcomes, it is necessary to understand whether these influences are general to self-reporting of traumatic experiences, or whether they are specific to retrospectively self-reported childhood trauma. Retrospective reporting of trauma that occurred in childhood may be more likely to be affected by these individual differences than more recent traumatic events, for example those occurring in adulthood ^33^.

Due to the large period of time between the occurrence of the experiences and the recall, memories of early experiences could be more easily forgotten or modified relative to memories of recent events. For example, traumatic childhood memories may be suppressed by competing later memories ^34^, amplified by consistent subsequent events, or influenced by subsequent discussion or feedback, such as in psychotherapy ^11^. Memories may not be encoded or consolidated, for example, if the event was not considered distressing at the time ^6^. Furthermore, most individuals are unable to recall even salient experiences from the first few years of life due to infantile amnesia ^35^. The susceptibility of childhood memories to these biases in recall may in part account for the inconsistency in retrospective reports of adverse childhood experiences over time ^36^. If these factors do impact retrospective self-reporting of childhood experiences, we would expect that measures of contemporaneous adulthood traumas may not be associated with the same characteristics.

In this study, we aimed to increase understanding of the pathways through which self-reported trauma acts as a risk factor for poor outcomes. The primary aim was to investigate reporter characteristics associated with retrospective self-reports of childhood trauma, using polygenic scores of cognitive, personality and psychopathological traits. To assess the presence of rGE, we investigated whether these associations remain after controlling for environmental adversity across development. As a secondary aim, we assessed the specificity of these findings to retrospectively self-reported childhood trauma, by comparing associations with self-reports of contemporaneous traumas in adulthood, specifically partner abuse and major life events.

## Methods

### Sample characteristics

The data used were from the Twins Early Development Study (TEDS) ^37^. TEDS is a longitudinal study of over 15,000 twin pairs born in England and Wales between 1994-1996, identified through birth records. At 13 timepoints between birth and age 21, TEDS has collected data on the home environment, physical and mental health, personality and cognitive abilities. The study sample included a subset of unrelated TEDS twins who had both genetic data and data on self-reported trauma at age 21 (n = 3963).

Ethical approval for TEDS has been provided by the King’s College London Ethics Committee (reference: PNM/09/10–104). Written informed consent was obtained from parents during childhood and from twins themselves from age 16 onwards prior to each wave of data collection.

The demographic characteristics of the study sample are shown in Table 3. The majority of the study participants were female (62%), dizygotic twins (62%) of white race (99%). The study sample is fairly representative of the full TEDS cohort and the UK population on family socioeconomic measures. The increased proportion of females in the study sample reflects the gradual decrease in male participants in recent waves of TEDS data collection ^37,38^. At first contact, participants’ race was reported by parents, from the categories of ‘Asian’, ‘Black’, ‘Mixed race’, ‘White’ and ‘Other’. We acknowledge that these categories are limited for representing participant identity. The high proportion of participants identified as white race corresponds to all genotyped participants being of European ancestry.

**Table 3.** Demographic characteristics of the Twins Early Development Study (TEDS) study sample (n = 3963), as compared to the full TEDS sample and UK Census

|  | Number of families | % White* | % Mothers with A- levels or higher | % Mother employed | % Father employed | Female (%) | MZ (%) |
| --- | --- | --- | --- | --- | --- | --- | --- |
| TEDS study sample | 3963 | 99% | 44% | 48% | 89% | 62% | 38% |
| TEDS full cohort | 13722 | 92% | 36% | 43% | 92% | 50% | 33% |
| UK Census | 8221 | 93% | 32% | 49% | 89% | 51% | 34% |

Note. UK data from the 2000 General Household Survey <sup>39</sup> are used rather than more recent data because they provide more appropriate comparisons for TEDS twins who were born 1994-96. The % MZ data represents the average % MZ in England and Wales from 1994-6 <sup>40</sup>. A-levels are the national educational exam taken at 18 years of age in the UK. MZ=monozygotic twins.
\*We acknowledge that these data are limited for representing participant identity, as discussed in text

### Measures

#### Childhood trauma

The outcome measures in these analyses were all from the TEDS-21 wave of data collection, when the twins’ ages ranged between 21 and 25 years ^38^. The primary outcome measure was retrospectively self-reported childhood trauma, measured using eight items assessing emotional and physical abuse, derived from the Avon Longitudinal Study of Parents and Children “Life at 22+” questionnaire ^41^ (Supplementary Table 1). Participants reported how frequently they experienced these types of abuse during their childhood, on a scale of ‘Never’ (0) to ‘Very often’ (4), with total scores ranging from 0 to 32. The average total score in the study sample was 5.2 (SD = 4.5).

#### Adult trauma

As a planned control, we compared findings for childhood trauma with results based on two measures of recent adult trauma: partner abuse and major life events. Self-reported partner abuse, including physical abuse, emotional abuse and control was assessed using six items adapted from the ‘Intimate Partner Violence Questions’ section of the Centers for Disease Control and Prevention Violence Prevention questionnaire ^42^ (Supplementary Table 1). Participants reported the extent to which they agreed with six statements describing their current or past partner on a scale of ‘Strongly disagree’ (0) to ‘Strongly agree’ (4), with total scores ranging from 0 to 24. The average total score in the study sample was 4.5 (SD = 5.8). Self-reported major life events occurring since the age of 16 were assessed using 11 items adapted from the ‘negative’ life events items of the Coddington scale, including becoming homeless or seriously ill ^43^ (Supplementary Table 1). Participants were asked to report whether each event had occurred since the age of 16 and the extent to which it affected them, on a scale of ‘No, did not happen’ (0) to ‘Yes, affected me a lot’ (4), with total scores ranging from 0 to 44. The average total score in the study sample was 3.3 (SD = 3.7).

#### Genome-wide polygenic scores

Genome-wide polygenic scores for 21 relevant psychiatric, cognitive, anthropometric and personality traits derived from the largest genome-wide association studies (GWAS) were selected (Supplementary Table 2). The majority of these polygenic scores had been previously constructed in TEDS for use in the prediction of cognitive and behavioural traits ^44–46^. In cases where TEDS participants formed part of the discovery sample, polygenic scores were created from GWAS with these individuals excluded. DNA samples obtained from saliva and buccal cheek swabs were genotyped on either Affymetrix GeneChip 6.0 or Illumina HumanOmniExpressExome-8v1.2 arrays. Data from both platforms underwent common quality control procedures. Individuals were removed based on call rate (<0.98), suspected non-European ancestry, heterozygosity, and relatedness other than twin status. Single-nucleotide polymorphisms (SNPs) SNPs were excluded if the minor allele frequency was smaller than 0.5%, if more than 2% of genotype data were missing, or if the Hardy Weinberg p-value was lower than 10^−5^. Full genotyping and quality control processes as detailed elsewhere ^47^ are provided in the Supplementary Methods. Polygenic scores were constructed using LDpred ^48^, with the proportion of genetic markers assumed to be contributing to the trait set to 1 ^49^.

#### Environmental measures

Environmental factors capturing environmental adversity across development were selected, including maternal depression ^50^, economic disadvantage ^51^, family cohesion ^52^, parental feelings or discipline ^12^, negative life events ^12^ and peer difficulties ^53^. These measures were selected as they have previously been associated with retrospectively self-reported childhood trauma, and indicate the experience of challenging early environments that tend to co-occur with childhood trauma.

Measures of early environmental adversity and ages collected are given in Supplementary Table 3. Early adversities were separated according to whether they were parent- or self-reported. We highlight this distinction because reports by parents and children typically show poor agreement, partly because they provide different perspectives on the family environment ^11,54^. As the nature of rGE effects vary across development, environmental adversities in pre-school (age 1-4), middle childhood (7-10) and early adolescence (12-16) were summed to create five composite scores of parent- and self-reported environmental adversity in each developmental period. Participants were included providing that they had data for more than 75% of variables, those missing more than 25% of variables at each age were excluded. The collection of self-reported measures began when the twins were aged 9, resulting in five composite scores of early environmental adversity: parent-reported adversities in pre-school, middle childhood and early adolescence, and self-reported early environmental adversities in middle childhood and early adolescence.

### Data handling

All predictors were standardised by centring and dividing by two standard deviations, resulting in a mean of 0 and a standard deviation of 0.5, using the rescale function from the R package ‘arm’ ^55^.

To account for participants with missing environmental adversity composite scores, multiple imputation was conducted using the R package ‘mice’, using a prediction matrix containing the five composite scores, polygenic scores, outcomes and covariates ^56,57^. The average missingness in the sample was 3.6%, the number of imputed datasets was 25, and the number of iterations was 20 ^58^. All analyses were then performed through ‘mice’ with the imputed data sets.

### Analysis

The primary aim of this study was to investigate whether genetic liabilities for psychiatric, cognitive, anthropometric and personality traits are associated with retrospective self-reports of childhood trauma, and the extent to which these associations are driven by gene-environment correlation. We addressed this aim using three analyses.

First, we assessed whether polygenic scores were associated with self-reported childhood trauma. Univariable linear regression models were run to assess the association between each polygenic score and retrospectively self-reported childhood trauma. Subsequently, a multivariable linear regression model was run, including all 21 polygenic scores.

Second, we tested the assumption that the composite scores of environmental adversity across development were associated with self-reported childhood trauma, as predicted from previous literature. Univariable linear regression models were run to assess the associations between each of the five environmental composite scores and retrospectively self-reported childhood trauma. Then a multivariable linear regression model was run, including all five environmental composite scores.

Finally, to assess the presence of rGE, we investigated whether the associations between polygenic scores and self-reported childhood trauma remained after controlling for environmental adversity across development. To do so, we regressed retrospectively self-reported childhood trauma on both the polygenic scores and the five composite scores of environmental adversity across development in one multivariable linear regression model.

As a secondary aim, we assessed the specificity of these findings to retrospectively self-reported childhood trauma, by comparing associations with self-reports of contemporaneous traumas in adulthood. To do so, we repeated the third step of the analysis for the two secondary outcomes, regressing self-reported partner abuse and major life events since age 16 on the polygenic scores and environmental composite scores, in two multivariable linear regression models.

All analyses were conducted using the lm function within ‘mice’, in R version 4.0.3 ^59^.

### Covariates and multiple testing

In all analyses, birth year (cohort), sex, and genotyping batch were included as covariates. In analyses of polygenic scores, the first 10 genetic principal components were also included. Corrections for multiple testing were applied using the Benjamini Hochberg false discovery rate (FDR) adjustment, with a threshold of 0.05, which controls for the expected proportion of false positives among the significant results ^60^.

## Results

### Association between polygenic scores and environmental adversity across development with adulthood retrospective self-reports of childhood trauma

#### Association between polygenic scores and retrospective self-reports of childhood trauma

Figure 1 shows the association between genome-wide polygenic scores and retrospectively self-reported childhood trauma at age 21, derived from univariable and multivariable linear regression models. Effect sizes (β) with 95% confidence intervals (CIs) are shown for a two standard deviation increase in polygenic scores.

**Figure 1.**
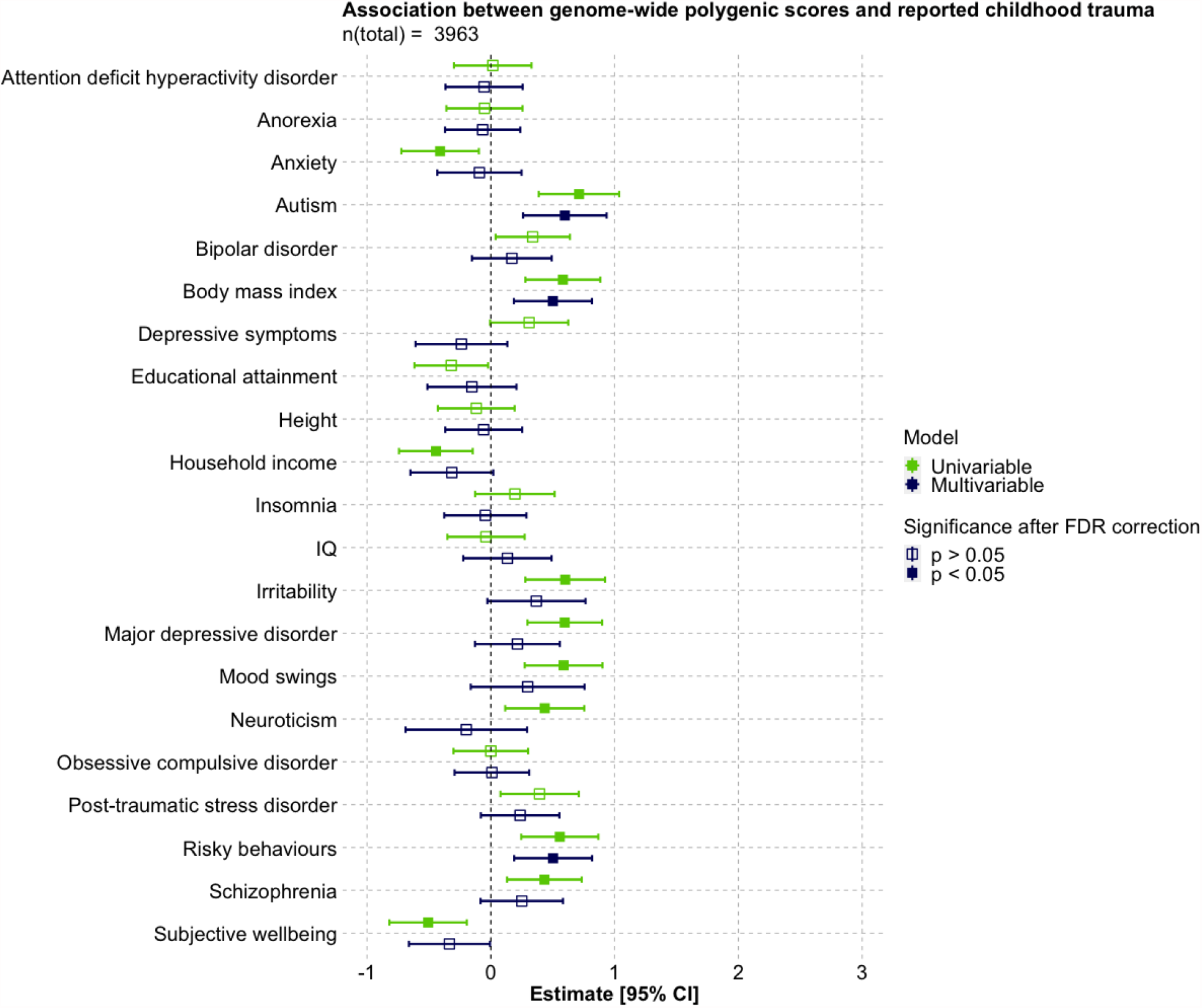
Association between genome-wide polygenic scores and retrospectively self-reported childhood trauma at age 21, derived from univariable and multivariable linear regression models (n = 3963).

In the univariable linear regression models, polygenic scores for ASD, BMI, irritability, major depressive disorder (MDD), mood swings, neuroticism, risky behaviours and schizophrenia were positively associated with retrospectively self-reported childhood trauma after FDR correction for multiple testing. A two standard deviation increase in these polygenic scores was associated with an increase in self-reported childhood trauma ranging from β = 0.43 (95% CI 0.13, 0.73) for schizophrenia to β = 0.71 (95% CI 0.39, 1.04) for ASD. Polygenic scores for anxiety (β = -0.41; 95% CI -0.72, -0.10), household income (β = -0.44; 95% CI -0.74, -0.15) and subjective well-being (β = -0.51; 95% CI -0.82, -0.19) were negatively associated with self-reported childhood trauma. When all polygenic scores were included in a multivariable linear regression model, only scores for ASD (β = 0.60; 95% CI 0.26 - 0.94), BMI (β = 0.50; 95% CI 0.19, 0.82) and risky behaviours (β = 0.50; 95% CI 0.19, 0.82) remained independently associated with retrospectively self-reported childhood trauma after FDR correction for multiple testing.

#### Association between environmental adversity across development and retrospective self-reports of childhood trauma

Figure 2 shows the associations between the five composite scores of parent-and self-reported environmental adversity in pre-school, middle childhood and early adolescence with retrospectively self-reported childhood trauma at age 21, derived from univariable and multivariable linear regression models. Effect sizes (β) with 95% CIs are shown for a two standard deviation increase in composite scores.

**Figure 2.**
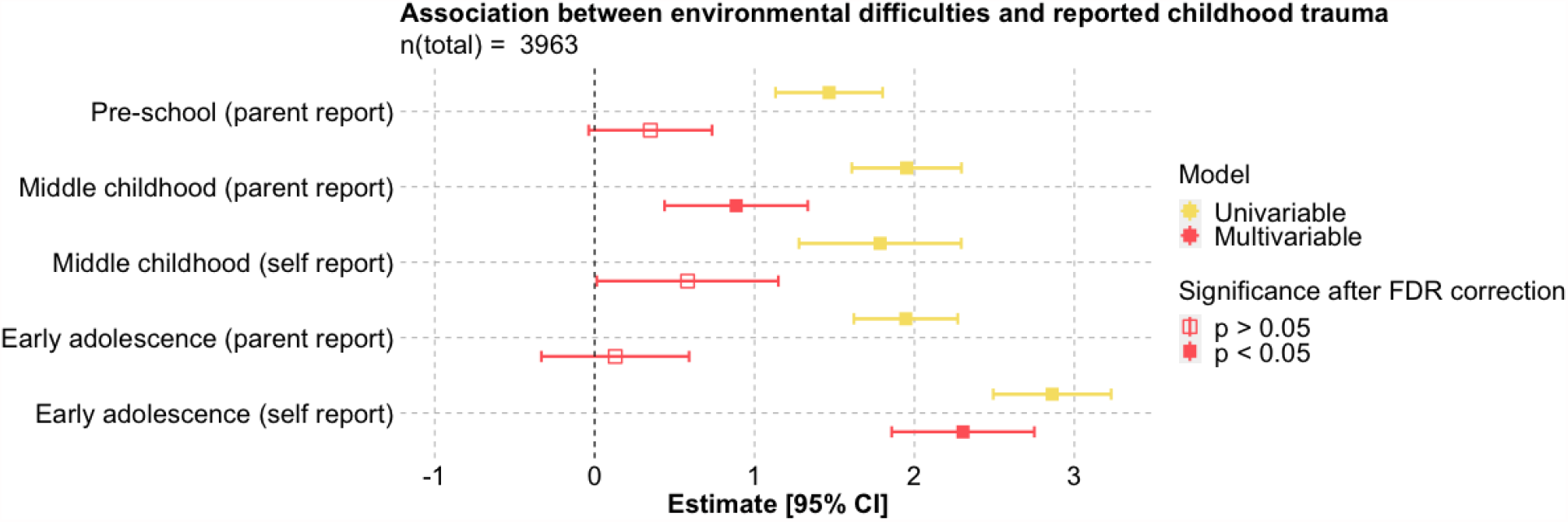
Association between composite scores of environmental adversity and retrospectively self-reported childhood trauma at age 21, derived from univariable and multivariable linear regression models (n = 3963).

As anticipated, in univariable linear regression models all five composite scores of environmental adversity were associated with retrospectively self-reported childhood trauma after FDR correction for multiple testing. A two standard deviation increase in any composite score was associated with an increase in self-reported childhood trauma ranging from β = 1.47 (95% CI 1.13, 1.80) for parent-reported environmental adversity in pre-school to β = 2.86 (95% CI 2.49, 3.23) for self-reported environmental adversity in early adolescence. When all composites were included in a multivariable linear regression model, only parent-reported environmental adversity in middle childhood (β = 0.89; 95% CI 0.44, 1.33) and self-reported environmental adversity in early adolescence (β = 2.30; 95% CI 1.86, 2.75) remained independently associated with retrospectively self-reported childhood trauma after FDR correction for multiple testing.

#### Independent associations between polygenic scores and environmental adversity across development with retrospective self-reports of childhood trauma

Figure 3 shows the associations between the polygenic scores and environmental adversity composite scores with retrospectively self-reported childhood trauma at age 21, derived from the multivariable linear regression model. Effect sizes (β) with 95% CIs are shown for a two standard deviation increase in composite scores and polygenic scores.

**Figure 3.**
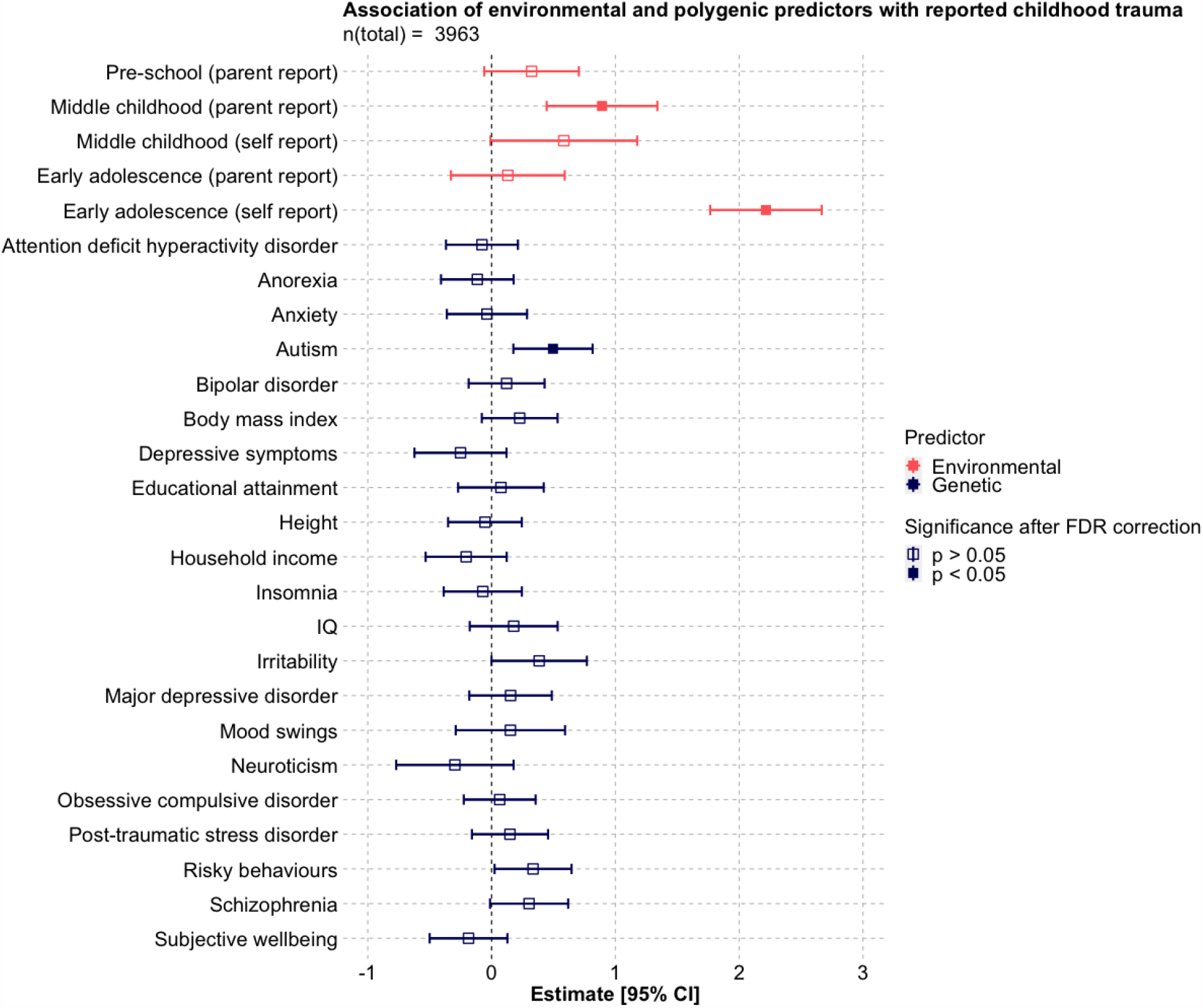
Association between composite scores of environmental adversity and genome-wide polygenic scores with retrospectively self-reported childhood trauma at age 21, derived from a multivariable linear regression model (n = 3963).

In the model of environmental composite scores and polygenic scores, only parent-reported environmental adversity in middle childhood (β = 0.89; 95% CI 0.44, 1.34), self-reported environmental adversity in early adolescence (β = 2.22; 95% CI 1.77, 2.67) and the polygenic score for ASD (β = 0.50; 95% CI 0.18, 0.82) remained independently associated with retrospectively self-reported childhood trauma.

### Comparison to self-reports of contemporaneous adulthood trauma

Post-hoc analyses revealed that three measures of self-reported trauma at age 21 were all weak to moderately correlated. The strongest correlation was between partner abuse and life events (Pearson’s *r* = 0.31), followed by childhood trauma and life events (*r* = 0.30), then childhood trauma and partner abuse (*r* =0.25).

Figures 4 and 5 show the associations between environmental composite scores and polygenic scores, with self-reported partner abuse and major life events at age 21, derived from multivariable linear regression models. Effect sizes (β) with 95% CIs are shown for a two standard deviation increase in composite scores and polygenic scores.

**Figure 4.**
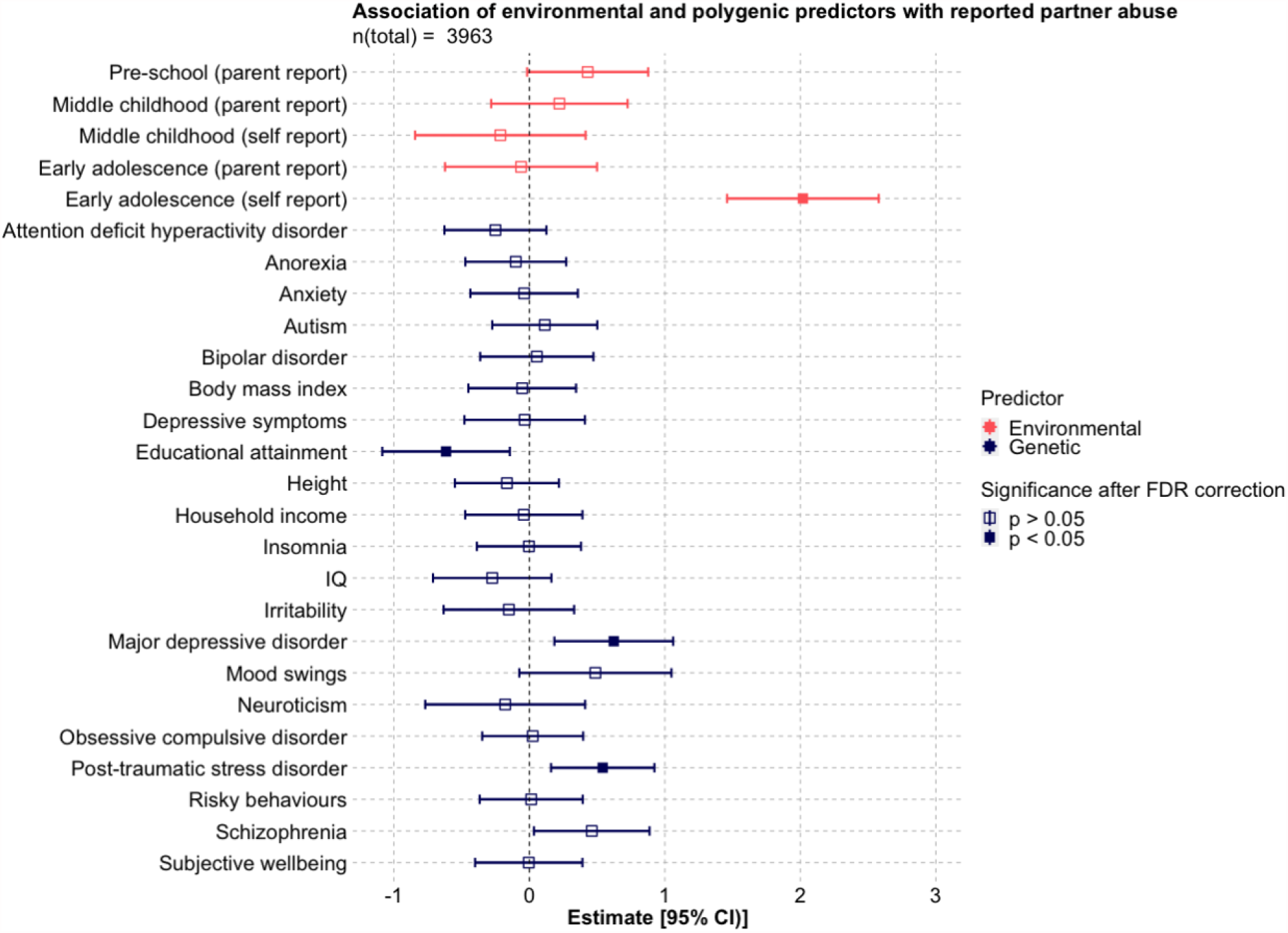
Association between environmental adversity and polygenic scores with self-reported partner abuse at age 21, derived from a multivariable linear regression model (n = 3963).

**Figure 5.**
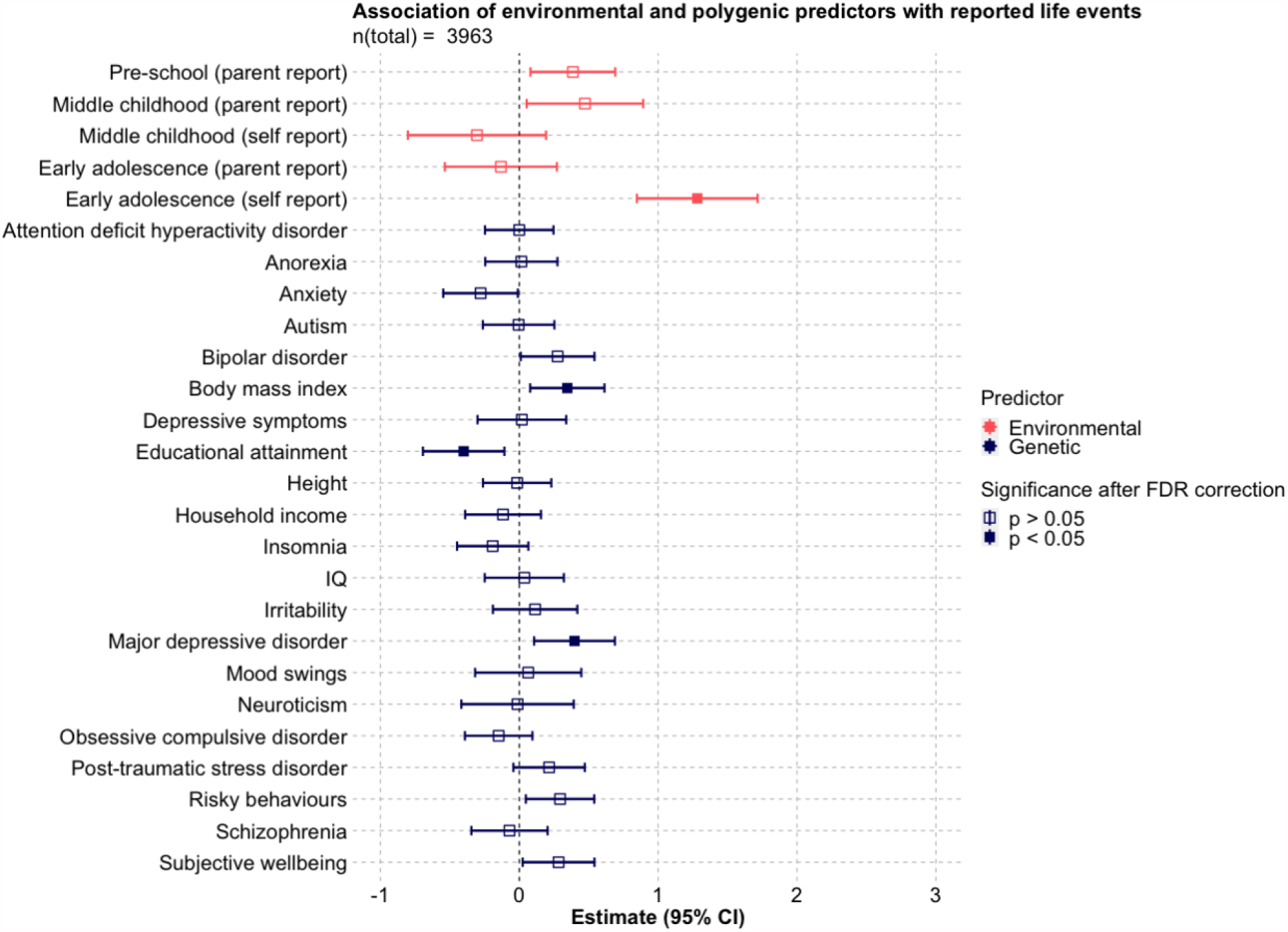
Association between environmental adversity and polygenic scores with self-reported major life events at age 21, derived from a multivariable linear regression model (n = 3963).

Self-reported environmental adversity in early adolescence, and the polygenic scores for MDD and educational attainment were associated with both self-reported partner abuse (early adolescence environmental adversity β = 2.02; 95% CI 1.46, 2.58; MDD β = 0.62; 95% CI 0.18, 1.06; educational attainment β = -0.61; 95% CI -1.09, -0.14) and major life events (early adolescence environmental adversity β = 1.28; 95% CI 0.85, 1.72; MDD β = 0.40; 95% CI 0.11, 0.69; educational attainment β = - 0.40; 95% CI -0.69, -0.11). In addition, the polygenic score for post-traumatic stress disorder (PTSD) was associated with partner abuse (β = 0.54; 95% CI 0.16, 0.92) and the polygenic score for BMI was associated with self-reported major life events (β = 0.35; 95% CI 0.08, 0.61).

## Discussion

In this study we examined polygenic scores capturing genetic liability to cognitive, personality and psychopathological traits associated with retrospective self-reports of childhood trauma and whether these associations remain after controlling for environmental adversity across development. In multivariable analysis of polygenic scores, retrospectively self-reported childhood trauma was independently associated with polygenic scores for ASD, BMI and risky behaviours. In a multivariable analysis of environmental adversity composite scores, retrospectively self-reported childhood trauma was associated with parent-reported environmental adversity in middle childhood and self-reported environmental adversity in early adolescence. When both polygenic scores and environmental adversity composites were included together in one multivariable model, only associations with parent-reported environmental adversity in middle childhood, self-reported environmental adversity in early adolescence and the polygenic score for ASD remained significant. A different pattern of association was seen for contemporaneous self-reports of adult trauma. Environmental adversity self-reported in early adolescence and polygenic scores for major depressive disorder and educational attainment were associated with both self-reported partner abuse and major life events. In addition, the polygenic score for PTSD was associated with self-reported partner abuse, and the polygenic score for BMI was positively associated with self-reported major life events.

The robust association between retrospectively self-reported childhood trauma and the polygenic score for ASD replicates findings from a cohort of female nurses ^27^ and from the UK Biobank ^61^. There are two possible explanations for this finding. One explanation is gene-environment correlation, which occurs when genetic factors influence both an individual’s liability for a trait and the environments that they are exposed to ^28^. Increased genetic liability for autistic traits may lead to difficulty with social interaction, which could evoke adverse reactions, such as neglect or abuse from others (evocative gene-environment correlation), or increase the likelihood of being in dangerous environments (active gene-environment correlation) ^61^. Consistent with this explanation, another study found over-transmission of genetic risk for reporting childhood trauma from parents to autistic children but not to their non-autistic siblings ^14^. This suggests that the increased genetic risk of reporting childhood maltreatment in individuals with ASD may be partly explained by increased active and evocative gene-environment correlation, rather than passive. A second explanation of these findings is that individuals with high genetic risk for ASD may be more sensitive to experiencing, interpreting and/or reporting an event as traumatic ^61^. This explanation is more in line with the results of the present study, as the association between the polygenic score for ASD and retrospectively self-reported childhood trauma remained significant after controlling for environmental adversity in childhood and adolescence. Consistent with this explanation, the polygenic score for ASD was not found to be associated with prospectively measured parent-reported childhood trauma ^62^. Indeed, adults with ASD report a broad range of life events as being traumatic ^63^. Furthermore, they are at increased risk of developing a trauma-related disorder, even following events that would not meet the Diagnostic and Statistical Manual of Mental Disorders ^64^ criteria for trauma ^63^. The ASD polygenic score has also been associated with lower likelihood of responding to cognitive behavioural therapy (CBT) for depression, which may indicate that CBT does not target the main causes of depression-related distress in individuals with a higher genetic predisposition to ASD ^65^. This explanation does not imply that these events were not traumatic, as trauma is not only the nature of an event but also how it is experienced by the individual ^61^. Rather, it provides a framework for understanding how the subjective experience relates to the outcomes of reported trauma.

Our finding that polygenic scores for BMI and risky behaviours were no longer significant once controlling for environmental adversity likely reflects overlap in some of the mechanisms that these variables capture. The polygenic score for risky behaviours was derived from the first principle component of behaviours across the four domains of automobile speeding propensity, number of alcoholic drinks consumed per week, whether one has ever been a smoker, and number of lifetime sexual partners, capturing a general tendency towards risk-taking ^66^. Risk-taking behaviours and high BMI are anticipated to share behavioural pathways, due to common associations with impulsivity, self-control and reward-seeking behaviours ^67–69^. These results suggest that gene-environment correlation may account for some of the association between polygenic scores for these traits and retrospectively self-reported childhood trauma. This could be through passive gene-environment correlation, for example, children in households where parents engage in more risky behaviours, such as alcohol abuse, are both more likely to experience childhood adversity ^70^ and also inherit a predisposition to these behaviours. These associations may also be influenced by evocative gene-environment correlation, which refers to an individual’s genetically-influenced behaviours instigating certain responses from others in their environment. For example, impulsive behaviour in children is associated with negative parenting and harsh discipline strategies ^71,72^. In line with these mechanisms, our findings suggest the effects of genotype on the likelihood of self-reporting trauma are not solely direct, but they can also be environmentally mediated.

The associations between retrospectively self-reported childhood trauma and genetic risk for ASD, BMI and risky behaviours reflect the previously identified shared genetic architecture of these traits ^20,69,73^. However, unlike previous research, we did not find associations between retrospectively self-reported childhood trauma and polygenic risk for other psychopathological and personality traits. In a cohort of female nurses, in addition to the polygenic score for ASD, associations were found between self-reported childhood trauma and polygenic scores for major depressive disorder, bipolar disorder, neuroticism, schizophrenia and ADHD ^27^. A likely reason for this inconsistency is that the larger sample size in the nurse study (n = 11,315) resulted in higher power to detect small effects (OR range 1.05 -1.19). Nevertheless, in the nurses study, when analyses were conducted using polygenic scores created with only variants uniquely associated with each trait, the score most strongly associated with reported childhood trauma was that of ASD ^27^. Therefore, both studies indicate a robust association between genetic liability for ASD and retrospectively self-reported childhood trauma, independent of genetic liability for other disorders.

Our finding that retrospectively self-reported childhood trauma was associated with prospectively collected measures of early environmental adversity is consistent with previous literature showing that reports of abuse and neglect are more common in households experiencing financial difficulties ^51^, poor parental mental health ^50^, low parental involvement or warmth ^12^ and family disruption ^52^. We found that adversities in middle childhood were most strongly associated with retrospectively self-reported childhood trauma when reported by parents, whereas adversities in early adolescence were most strongly associated when self-reported by the child. This may be due to differences in the experiences assessed in the parent- and child-reported measures. However, it is also consistent with research indicating that parents are more reliable reporters of very personal experiences in younger children, whereas for children older than 12 self-reports are more reliable, particularly for negative events ^74,75^. Furthermore, the fact that retrospective self-reports of trauma are associated with parent reports of environmental adversity in middle childhood, as well as self-reported environmental adversity, suggests that retrospective self-reports are not only associated with subjective experiences but also broader, more objective, environmental adversity.

When exploring correlates of self-reported contemporaneous traumas in adulthood, a different pattern of results was found. Polygenic scores for educational attainment and major depressive disorder were also associated with both self-reported partner abuse and major life events, while the polygenic score for PTSD was associated with self-reported partner abuse, and the polygenic score for BMI was associated with self-reported major life events. For major depressive disorder, this is in line with previous research showing that genetic liability for major depression may contribute to the subjective experience, interpretation or reporting of traumatic life events ^76,77^. Gene-environment correlation is also likely to explain part of this association, whereby those at greater genetic risk for depression are more likely to experience their environment as being aversive ^78^. The association between the polygenic score for depression and self-reported adult traumas, but not childhood trauma, is consistent with the hypothesis of gene-environment correlation for depression increasing throughout adolescence into adulthood ^24^.

Similarly, the types of cognitive and non-cognitive traits, such as motivation and intellectual interest, that reflect genetic liability for educational attainment may themselves influence the environments that individuals experience ^79,80^. Genetic variants associated with traits such as educational attainment and BMI could also reflect genetic influences on related environmental risks, such as economic disadvantage ^81^. As environmental adversity in late adolescence and early adulthood was not included in these analyses, it cannot be inferred whether genetic predisposition to these traits is associated with self-reports of adult trauma directly, or whether these associations are environmentally-mediated. However, what these results do indicate is that genetic liability to these psychiatric and sociodemographic factors may play a greater role in the self-reporting of contemporaneous adult traumas than in retrospective reports of childhood trauma.

One limitation of this study was the use of composite scores, comprising the total environmental adversity in pre-school, middle childhood and early adolescence. Using composite scores means it is difficult to infer which specific environmental adversities within each developmental period are driving associations with self-reported trauma. Furthermore, not all types of environmental adversity were included at each time point. For example, relationship problems were only assessed in middle childhood. Therefore, associations specific to a particular developmental stage could be confounded by the types of adversity assessed at that time. However, this approach enabled the inclusion of a broader scope of environmental adversities across development, from birth to age 16. Additionally, it allowed for the separation of parent-reported and self-reported environmental experiences, enabling the evaluation of common rater bias ^82^. Another limitation of this analysis was the conceptualisation of childhood trauma as only physical or emotional abuse. Childhood trauma can take several other forms, including physical and emotional *neglect* and sexual abuse, which are associated with differential reporting patterns, heritability estimates and outcomes ^8,13,27^. Nevertheless, different types of childhood trauma are found to have substantial genetic overlap ^14^. Finally, despite being broadly representative of the UK population, the TEDS cohort shows some bias in sociodemographic characteristics ^37^. Of particular note, only participants of European ancestry have been included in the genotyped subset. This is due to many current polygenic scores having poor predictive performance in individuals from more diverse ancestry groups, as the GWAS that these scores are derived from have only included individuals of European ancestry in order to increase homogeneity. The need for large-scale GWAS in diverse human populations, and subsequently polygenic scores with predictive power across ancestral groups, is an immediate priority for research in this field ^83^.

These findings have several implications. First, they indicate that self-reports of trauma are associated with heritable characteristics of the reporter. This reinforces the importance of using genetically sensitive designs when assessing associations between self-reported experiences and later life outcomes ^84,85^. Research that does not account for the confounding role of genetics should take particular care not to draw causal inferences. In contrast, genetically sensitive designs can help disentangle the complex mechanisms through which self-reports of childhood trauma are linked with poor outcomes, informing preventative and therapeutic strategies. Hence, there is benefit in not focusing on genetic or environmental correlates in isolation, but instead conducting comprehensive research that reflects the complexity of the underlying processes. These findings also suggest that the genetic predispositions that confound associations between self-reports of trauma and poor outcomes differ for childhood and adult traumas. Finally, it is important to stress that these results show only associations with self-reported experiences and do not imply that genetic predisposition for these traits would cause a person to experience adversity.

## Conclusions

Self-reports of trauma are associated with genetically-influenced reporter characteristics. Retrospectively self-reported childhood trauma and self-reports of contemporaneous adult traumas display different patterns of association. Studies of the association between retrospectively self-reported childhood trauma and later life outcomes should consider that genetically-influenced reporter characteristics may confound these relationships, both directly and through gene-environment correlation.

## Data Availability

The data that support the findings of this study came from the Twins Early Development Study (TEDS). Eligible researchers can apply for access to the TEDS data: https://www.teds.ac.uk/researchers/teds-data-access-policy.

## Acknowledgements

We gratefully acknowledge the ongoing contribution of the participants in the Twins Early Development Study (TEDS) and their families.

