## Supplementary Methods for "Genetic and early environmental predictors of adulthood self-reports of trauma"

**Genotyping and Quality Control information (Selzam, S. *et al.* Evidence for gene-environment correlation in child feeding: Links between common genetic variation for BMI in children and parental feeding practices. *PLoS Genet.* 14, e1007757 (2018))**

DNA for 8,122 individuals (including 3,607 dizygotic co-twin samples) was extracted from saliva and buccal cheek swab samples and hybridized to HumanOmniExpressExome-8v1.2 genotyping arrays at the Institute of Psychiatry, Psychology and Neuroscience Genomics & Biomarker Core Facility. The raw image data from the array were normalized, pre-processed, and filtered in GenomeStudio according to Illumina Exome Chip SOP v1.4. (http://confluence.brc.iop.kcl.ac.uk:8090/display/PUB/Production+Version%3A+Illumina+Exome+Chip+SOP+v1.4). In addition, prior to genotype calling, 919 multi-mapping SNPs and 501 samples with call rate <0.95 were removed. The ZCALL program was used to augment the genotype calling for samples and SNPs that passed the initial QC.

DNA from 3,747 samples was extracted from buccal cheek swabs and genotyped at Affymetrix, Santa Clara, California, USA. From this sample, 3,665 samples were successfully hybridized to AffymetrixGeneChip 6.0 SNP genotyping arrays (http://www.affymetrix.com/support/technical/datasheets/genomewide_snp6_datasheet.pdf) using experimental protocols recommended by the manufacturer (Affymetrix Inc., Santa Clara, CA). The raw image data from the arrays were normalized and pre-processed at the Wellcome Trust Sanger Institute, Hinxton, UK for genotyping as part of the Wellcome Trust Case Control Consortium 2 (https://www.wtccc.org.uk/ccc2/) according to the manufacturer's guidelines (http://www.affymetrix.com/support/downloads/manuals/genomewidesnp6_manual.pdf). Genotypes for the Affymetrix arrays were called using CHIAMO (https://mathgen.stats.ox.ac.uk/genetics_software/chiamo/chiamo.html).

After initial quality control and genotype calling, the same quality control was performed on the samples genotyped on the Illumina and Affymetrix platforms separately using PLINK[1,2], R[3], BCFtools[4], and EIGENSOFT[5,6].

Samples were removed from subsequent analyses on the basis of call rate (<0.98), suspected non-European ancestry, heterozygosity, and relatedness other than dizygotic twin status. SNPs were excluded if the minor allele frequency was smaller than 0.5%, if more than 2% of genotype data were missing, or if the Hardy Weinberg $p$-value was lower than $10^{-5}$. Non-autosomal markers and indels were removed. Association between SNP and the platform, batch, plate or well on which samples were genotyped was calculated; SNPs with an effect $p$-value < $10^{-4}$ were excluded. A total sample of 10,346 samples (including 3,320 dizygotic twin pairs and 7,026 unrelated individuals), with 7,289 individuals and 559,772 SNPs genotyped on Illumina and 3,057 individuals and 635,269 SNPs genotyped on Affymetrix remained after quality control.

Genotypes from the two platforms were separately phased using EAGLE2[7], and imputed into the Haplotype Reference Consortium (release 1.1) using the Positional Burrows-Wheeler Transform method[8] through the Sanger Imputation Service[9]. Prior to merging, variants with info <0.75 were excluded and non-overlapping SNPs between platforms were removed. After merging, minor allele frequency differences between platforms were tested for and SNPs with an effect p-value < $10^{-4}$, and Hardy Weinberg p-value > $10^{-5}$ were removed. Using these criteria, 7,363,646 genotyped and well-imputed SNPs were retained for the analyses.

Principal component analysis was performed on a subset of 39,353 common (MAF > 5%), perfectly imputed (info = 1) autosomal SNPs, after stringent pruning to remove markers in linkage disequilibrium ($r^2$ > 0.1) and excluding high linkage disequilibrium genomic regions so as to ensure that only genome-wide effects were detected.
